# Artificial intelligence and MRI: the source of a new epilepsy taxonomy

**DOI:** 10.1101/2022.11.10.22282047

**Authors:** Fenglai Xiao, Lorenzo Caciagli, Britta Wandschneider, Daichi Sone, Alexandra L. Young, Sjoerd B. Vos, Gavin P. Winston, Yingying Zhang, Wenyu Liu, Dongmei An, Baris Kanber, Dong Zhou, Josemir W. Sander, John S. Duncan, Daniel C. Alexander, Marian Galovic, Matthias J. Koepp

## Abstract

Artificial intelligence (AI)-based tools are widely employed, but their use for diagnosis and prognosis of neurological disorders is still evolving. We capitalise on a large-scale, cross-sectional structural MRI dataset of 814 people with epilepsy. We use a recently developed machine-learning algorithm, Subtype and Stage Inference (SuStaIn), to develop a novel data-driven disease taxonomy based on distinct patterns of spatiotemporal progression of brain atrophy. We identify two subtypes common to focal and idiopathic generalised epilepsies, characterised by neocortical-driven or basal ganglia-driven progression, and a third subtype, only detected in focal epilepsies, characterised by hippocampus-driven progression. We corroborate external validity via an independent cohort of 254 people and decode associations between progression subtypes and clinical measures of epilepsy severity. Our findings suggest fundamental processes underlying the progression of epilepsy-related brain atrophy. We deliver a novel MRI- and AI-guided epilepsy taxonomy, which could be used for individualised prognostics and targeted therapeutics.

## Introduction

Epilepsy is a common neurological disorder, often chronic and disabling.^1^ The current classification of epilepsy types and syndromes is based on seizure semiology,^2,3^ aetiology,^4^ imaging,^5,6^ and other diagnostic data.^7–9^ It informs treatment and prognosis.^10^ Structural MRI provides reproducible quantitative measures of brain morphology,^11^ which can be conceptualised as biomarkers of pathological processes.^12,13^ In focal epilepsy, cortical thinning encompasses large-scale cortico-subcortical networks.^14^ In idiopathic generalized epilepsy (IGE), structural abnormalities predominantly involve thalamocortical circuitry,^15^ but also extend to fronto-temporo-parietal cortices.^16^ Recent evidence suggests that, in epilepsy, brain atrophy may progress over time.^17^ In cross-sectional studies, brain atrophy in focal epilepsy is related to disease duration, seizure frequency, and occurrence of focal-to-bilateral tonic-clonic seizures.^17^ Longitudinal studies highlight widespread cumulative grey matter atrophy, affecting regions beyond those near the seizure focus.^13^ In IGE, grey matter loss is more prominent with longer epilepsy duration and higher seizure frequency.^15,18,19^

While progressive brain atrophy may be a core imaging characteristic of epilepsy, the underlying neural processes remain poorly understood. Firstly, it is uncertain whether there is a consistent spatiotemporal sequence of atrophy progression over time. Secondly, it remains unclear whether trajectories may vary according to epilepsy types, e.g., focal and generalised epilepsies.^17^ Thirdly, the magnitude and extent of interindividual differences in progression paths and their relationship with clinical characteristics remain undetermined. Thus, there is a need to identify sources of interindividual variability to orient individual-tailored prognostics and treatment. Progress may be achieved by applying artificial intelligence, particularly techniques in the machine learning (ML) subfield, that are increasingly used in biomedical research.^20^ In epilepsy, imaging-based ML has successfully lateralised temporal lobe epilepsy (TLE),^21^ identified radiologically occult epileptogenic lesions,^22,23^ predicted epilepsy surgery outcomes,^24^ and typified individual-specific patterns of whole-brain structural reorganization relating to disease severity.^25^

Here we capitalise on an unsupervised, data-driven, hypothesis-free ML algorithm, *Subtype and Stage Inference* (SuStaIn),^26^ that was recently developed to capture patterns of disease progression in chronic conditions, particularly neurodegenerative disorders such as Alzheimer’s and Parkinson’s.^26–30^ SuStaIn reaches longitudinal inference from cross-sectional data. Specifically, it automatically identifies distinct spatiotemporal trajectories (patterns) of cumulative pathological alteration shown by measured biomarkers and quantifies their level of individual co-expression.^26,30,31^ Our study employs SuStaIn to decode individualised signatures of progressive cortico-subcortical atrophy in large focal and generalised epilepsy cohorts. We introduce a novel, ML-guided and MRI-based epilepsy taxonomy that combines categorical and dimensional perspectives by (i) quantifying main progression patterns of grey matter atrophy in each individual and (ii) identifying subgroups based on the dominant progression pattern. We also replicate our findings in an external validation cohort and relate ML-identified subtypes to clinical characteristics. We provide a novel stratification framework useful for individualized prognostics and targeted therapeutics by capturing fundamental processes underlying the progression of epilepsy-associated brain atrophy.

## Results

Our discovery cohort included 814 participants after data quality checks: (i) 503 with focal epilepsy (TLE: 328; frontal lobe epilepsy, FLE: 88; parietal lobe epilepsy, PLE: 27; occipital lobe epilepsy, OLE: 5; unclassified focal epilepsy: 55), of whom 418 with a lateralised seizure focus; (ii) 193 with IGE (juvenile myoclonic epilepsy, JME: 46; juvenile absence epilepsy, JAE: 43; IGE unclassified with generalised tonic-clonic seizures, GTCS, as primary seizure type: 104); and (iii) 118 healthy controls. All participants were assessed at UCL in London with a high-resolution T1-weighted MRI. After quality checks, the external validation cohort included 254 participants: (i) 122 with focal epilepsy (TLE/FLE/posterior quadrant: 50/50/22; 115 with a lateralised seizure focus); (ii) 61 with JME, and (iii) 71 healthy controls. All were assessed with a high-resolution T1-weighted MRI at West China Hospital of Sichuan University, Chengdu, China. Demographic and clinical characteristics are provided in Table 1. There were no significant differences in age at seizure onset between people with focal epilepsy and those with IGE in the discovery and validation cohorts (two-tailed, two-sample *t-*tests, discovery/validation; *t*_(181/630)_=-1.19/1.62, *p*=0.236/0.106).

**Table 1.** Demographic and clinical characteristics of discovery and external validation cohorts.

|  | **Discovery cohort** |  |  | **External validation cohort** |  |  |
| --- | --- | --- | --- | --- | --- | --- |
|  | *Focal epilepsy* | *IGE* | *Healthy Controls* | *Focal epilepsy* | *IGE* | *Healthy Controls* |
| **Participant number** | 503 | 193 | 118 | 122 | 61 | 71 |
| **Sex (F/M)** | 248/255 | 112/81 | 74/44 | 47/75 | 27/34 | 39/32 |
| **Age mean (SD)** | 35.0 (10.8) | 33.7 (11.4) | 36.3 (12.4) | 25.1 (8.6) | 20.6 (5.6) | 26.1 (6.9) |
| **Handedness** | R (433)/ L (63)/ A (7) | R (191)/ L (2) | R (108)/ L (10) | R (121)/ L (1) | R (61) | R (71) |
| **Age of onset mean (SD)** | 15.6 (12.2) | 13.9 (6.1)* | N/A | 15.2 (10.3) | 14.0 (3.0) | N/A |
| **Duration of epilepsy mean (SD)** | 19.9 (12.6) | 20.5 (12.2)* | N/A | 9.9 (7.2) | 5.0 (3.1) | N/A |
| **Lateralization of seizure focus (n)** | L (225)/ R (193)/ B (44)/ U (41) | N/A | N/A | L (83)/ R (32)/ B (4)/ U (3) | N/A | N/A |
| **Localization of seizure focus, or Epilepsy type (n)** | TLE (328)/ FLE (88)/ PLE (27)/ OLE (5)/ U (55) | JAE (43)/ JME (46)/ GTCS-unc (104) | N/A | TLE (50)/ FLE (50)/ PO (22) | JME (61) | N/A |
| **Localization of seizure focus of cases with proven lateralisation (n)** | TLE (281)/ FLE (81)/ PLE (24)/ OLE (4)/ U (28) | N/A | N/A | TLE (50)/ FLE (45)/ PO (20) | N/A | N/A |
| **Unilateral focal epilepsy (%)** | 418 (83.1%) | N/A | N/A | 115 (94.3%) | N/A | N/A |
| **† FBTCS (focal), GTCS (generalized), (% of patients)** | 59.6% | 53.9% | N/A | 62.4% | 35.9% | N/A |
| **HS (n)** | 96 | N/A | N/A | 34 | N/A | N/A |
| **Seizure frequency median (range)** | 3 (0-4) | 1 (0-4) | N/A | 2 (0-4) | 2 (0-4) | N/A |
| **ASM median (range)** | 2 (0-6) | 2 (0-4) | N/A | 2 (0-3) | 1 (0-2) | N/A |
| **Surgery n (%)** | 108 (21.5%) |  |  | 34 (27.9%) |  |  |

ASM: anti-seizure medication; F: female; FLE: frontal lobe epilepsy; GTCS: generalized tonic clonic seizure; GTCS-unc: IGE unclassified, with GTCS as primary seizure subtype; HC: healthy control; HS: hippocampal sclerosis; IGE: idiopathic generalized epilepsy; JAE: juvenile absence epilepsy; JME: juvenile myoclonic epilepsy; L: left; M: male; n: number; OLE:
occipital lobe epilepsy; PLE: parietal lobe epilepsy; PO: posterior quadrant; R: right; SD: standard deviation; TLE: temporal lobe epilepsy; U: undetermined. \*Reliable data pertaining to age at seizure onset and duration of epilepsy were available in 129 individuals with IGE of the discovery cohort. †Refers to at least one FBTCS (focal epilepsies) or GTCS (IGE) experienced in the year before the MRI.

We applied the SuStaIn algorithm (Fig. 1) to focal epilepsy and IGE samples separately to identify distinct spatiotemporal progression patterns of cortico-subcortical atrophy (Supplementary Fig. 1), termed *progression subtypes*. Each subtype is co-expressed to a different extent *in each participant* with epilepsy so that their within-individual sum amounts to 1. For categorical classification purposes, we then assigned each individual with epilepsy to their primary progression subtype based on the maximum likelihood of expression using a cut-off value of >50%, following prior work.^25,30^

**Figure 1.**
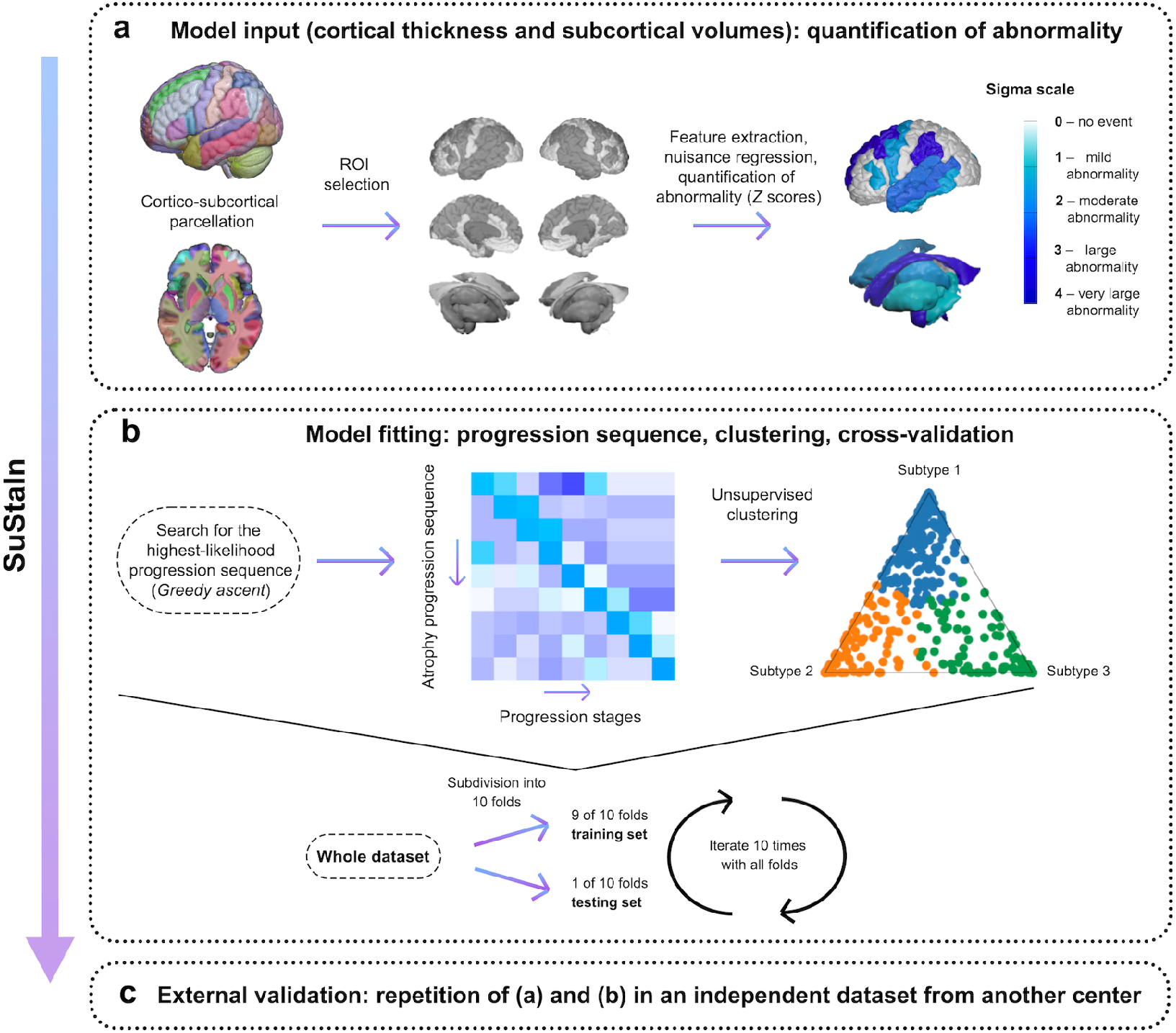
Visual schematic of the SuStaIn event-based model. We applied the SuStaIn algorithm to derive spatiotemporal patterns of progression of atrophy in large samples of people with focal epilepsy and IGE (total n>1000). The three main steps of the algorithm consist of: (a) *Model input*: selection of regions of interest, adjustment for nuisance variables, and conversion of regional grey matter metrics into z-scores relative to healthy control data; (b) *Model fitting*: computation of the best-fit probability distributions for normal and atrophic brain regions, identification of the most likely progression sequence, and quantification of uncertainty with cross-validation. An exemplary positional variance diagram, displayed on the left-hand side, shows an example of atrophy progression sequence with the highest likelihood on the *y-*axis, and the number of model stages (i.e., sequence positions) on the *x*-axis; the intensity of each entry corresponds to the proportion of Markov Chain Monte Carlo samples for which a certain region of the *y*-axis appears at the respective stage of the *x*-axis. An exemplary ternary plot shows the probability with which each individual is assigned to a given subtype, whereby each vertex represents the point at which membership of a given subtype is maximal (100%). The dots correspond to individual data and are labelled by final subtype classification: subtype 1 (blue); subtype 2 (orange) or subtype 3 (green). (c) *External validation*: repetition of procedures detailed in passages (a) and (b) for the external validation cohort, to corroborate reproducibility.

### SuStaIn identifies three focal epilepsy subtypes based on the spatiotemporal progression of atrophy

We identified three progression-based subtypes in focal epilepsy (Fig. 2a-d), each characterized by a sequence of *stages* (Fig. 2e): (1) a *cortical* progression subtype, dominant in 49.1% of cases [cross-validation folds: 0.85, 95% confidence interval (CI): 0.80-0.89; Fig. 2a], characterised by atrophy initially encompassing the superior and transverse temporal gyri and parietal operculum, followed by the superior frontal, middle frontal and precentral cortices, then by the precuneus and posterior cingulate cortex, and by subcortical areas only in late stages; (2) a *basal ganglia* subtype, dominant in 18.1% of cases (cross-validation folds: 0.78, 95% CI: 0.73-0.83; Fig. 2b), with initial involvement of the globus pallidus, followed by other basal ganglia regions, thalamus, and fronto-temporo-parietal cortices at later stages; and (3) a *hippocampal* subtype, dominant in 32.8% of cases (cross-validation folds: 0.88, 95% CI: 0.83-0.92; Fig. 2c), with a sequence first involving the hippocampus, followed by the thalamus, superior and middle temporal gyri, and then by other cortical areas.

**Figure 2.**
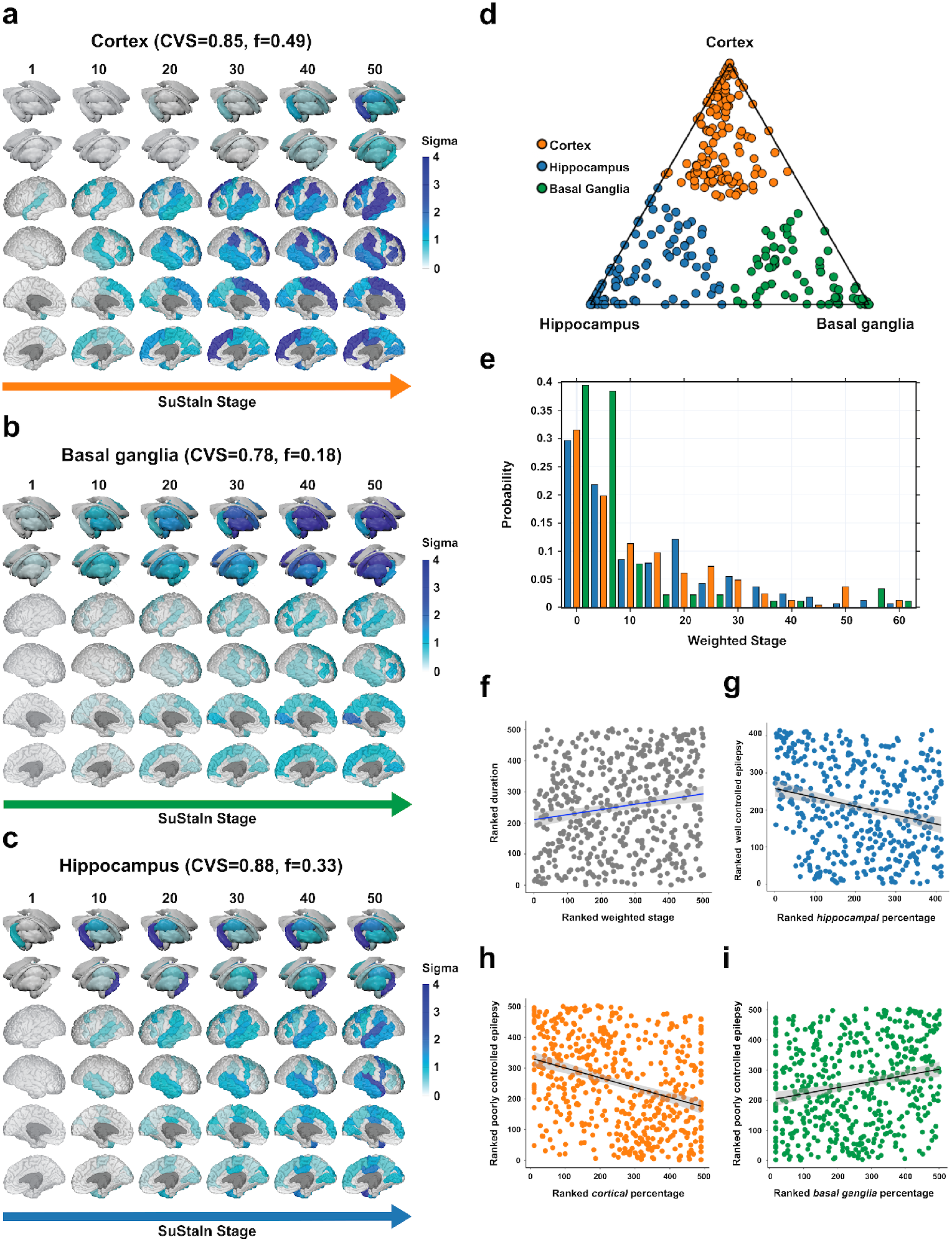
MRI-based progression subtypes in focal epilepsy. The figure shows the spatiotemporal patterns of progression of grey matter atrophy (subtypes; **a**. *cortical*; **b**. *basal ganglia*; **c**. *hippocampal*) identified via SuStaIn in the focal epilepsy discovery cohort. Each progression pattern in panels **a-c** consists of a sequence of stages with which cortical thickness and subcortical volumes reach different z-scores in people with epilepsy relative to healthy controls. The colour of each region indicates the severity of grey matter loss; white: unaffected areas; light blue: mildly affected areas (*z*-score=1-2); blue: moderately affected areas (*z*-score=2-3); and dark blue: severely affected areas (*z*-score >3); “CVS’’: cross-validation similarity. “*f*”: proportion of participants assigned to each subtype. Panel **d** shows the assignability of the disease subtype, operationalised as the distance from each vertex of the triangle, whereby each vertex represents the point at which membership of a given subtype is maximal (100%). Each participant was assigned to one subtype (orange, green, or blue) based on the maximum likelihood of subtype expression (cut-off value: >50%). Panel **e** shows the probability with which each participant from the focal epilepsy discovery cohort was assigned a specific SuStaIn stage (stage ranges: 0.002–62.424). Panel **f** shows the correlation between duration of epilepsy and weighted stage. Panel **g** shows a negative correlation between within-individual expression of *hippocampal* subtype and a marker of well controlled epilepsy [principal component (PC2); see main text]. Panels **h-i** show the correlations between within-individual expression of *cortical* and *basal ganglia* subtypes and a marker of poorly controlled epilepsy (PC1). Correlation analyses were conducted with Spearman’s *ρ*; the associated panels show ranked data.

In the external validation cohort, we also identified three subtypes with comparable progression patterns and proportion of people counted under each subtype: *cortical*: 41% (cross-validation folds: 0.82, 95% CI: 0.73-0.91); *hippocampal*, 37.7% (cross-validation folds: 0.81, 95% CI: 0.71-0.91); and *basal ganglia*: 21.3% (cross-validation folds: 0.90, 95% CI: 0.87-0.93). There were no significant differences in subtype prevalence between discovery and validation cohorts (χ^2^_2_=3.779, *p*=0.151), which corroborates the generalizability of our findings (Supplementary Fig. 2). Cross-validation analyses showed high reproducibility, with average similarity among cross-validation folds >78/>81% for each subtype in the discovery/validation cohorts. There were slight differences between discovery and validation cohorts: in the *cortical* subtype, the initial stages of the progression sequence were reversed, with temporal involvement following fronto-central involvement. For the *basal ganglia* subtype, the caudate was affected first, followed by the globus pallidus and the thalamus.

### Clinical characterisation of the focal epilepsy subtypes

We next characterised each subtype from a clinical standpoint (Table 2).

**Table 2.**
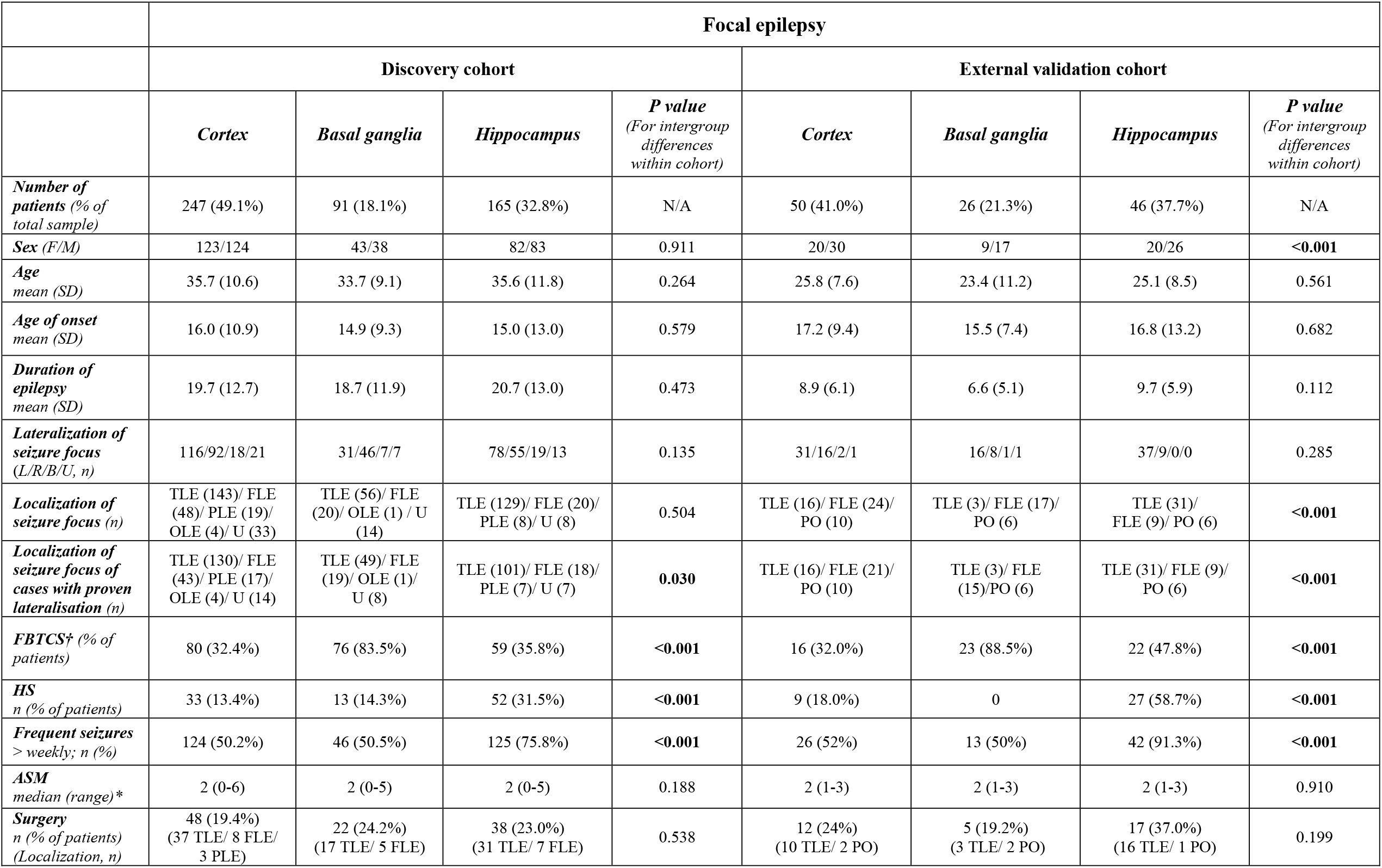
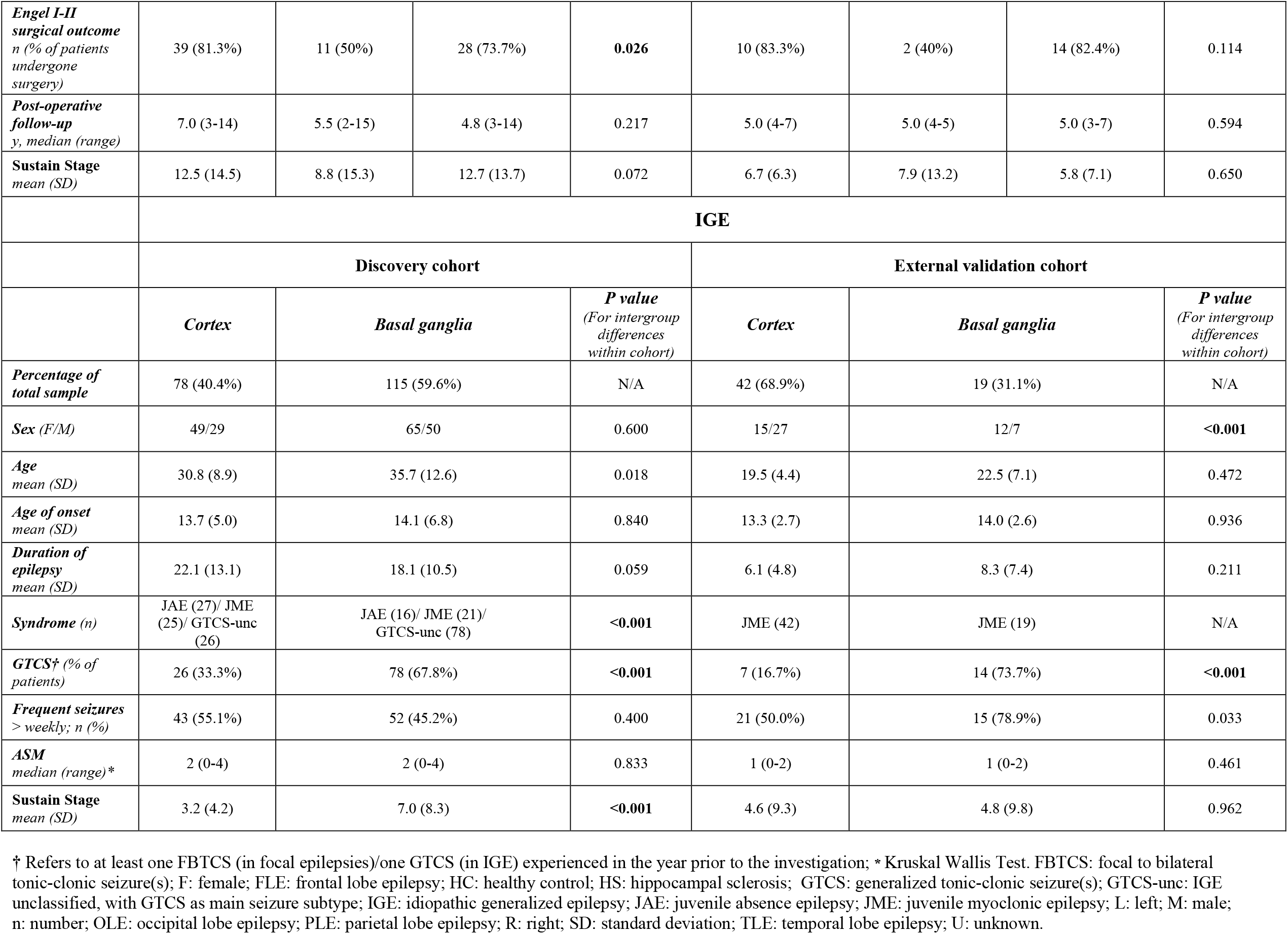
Characterisation of MRI-based focal epilepsy subtypes in the discovery and external validation cohorts.

|  | Focal epilepsy |  |  |  |  |  |  |  |
| --- | --- | --- | --- | --- | --- | --- | --- | --- |
|  | Discovery cohort |  |  |  | External validation cohort |  |  |  |
|  | *Cortex* | *Basal ganglia* | *Hippocampus* | *P value*<br>(For intergroup differences within cohort) | *Cortex* | *Basal ganglia* | *Hippocampus* | *P value*<br>(For intergroup differences within cohort) |
| *Number of patients (% of total sample)* | 247 (49.1%) | 91 (18.1%) | 165 (32.8%) | N/A | 50 (41.0%) | 26 (21.3%) | 46 (37.7%) | N/A |
| *Sex (F/M)* | 123/124 | 43/38 | 82/83 | 0.911 | 20/30 | 9/17 | 20/26 | **<0.001** |
| *Age mean (SD)* | 35.7 (10.6) | 33.7 (9.1) | 35.6 (11.8) | 0.264 | 25.8 (7.6) | 23.4 (11.2) | 25.1 (8.5) | 0.561 |
| *Age of onset mean (SD)* | 16.0 (10.9) | 14.9 (9.3) | 15.0 (13.0) | 0.579 | 17.2 (9.4) | 15.5 (7.4) | 16.8 (13.2) | 0.682 |
| *Duration of epilepsy mean (SD)* | 19.7 (12.7) | 18.7 (11.9) | 20.7 (13.0) | 0.473 | 8.9 (6.1) | 6.6 (5.1) | 9.7 (5.9) | 0.112 |
| *Lateralization of seizure focus (L/R/B/U, n)* | 116/92/18/21 | 31/46/7/7 | 78/55/19/13 | 0.135 | 31/16/2/1 | 16/8/1/1 | 37/9/0/0 | 0.285 |
| *Localization of seizure focus (n)* | TLE (143)/ FLE (48)/ PLE (19)/ OLE (4)/ U (33) | TLE (56)/ FLE (20)/ OLE (1) / U (14) | TLE (129)/ FLE (20)/ PLE (8)/ U (8) | 0.504 | TLE (16)/ FLE (24)/ PO (10) | TLE (3)/ FLE (17)/ PO (6) | TLE (31)/ FLE (9)/ PO (6) | **<0.001** |
| *Localization of seizure focus of cases with proven lateralisation (n)* | TLE (130)/ FLE (43)/ PLE (17)/ OLE (4)/ U (14) | TLE (49)/ FLE (19)/ OLE (1)/ U (8) | TLE (101)/ FLE (18)/ PLE (7)/ U (7) | **0.030** | TLE (16)/ FLE (21)/ PO (10) | TLE (3)/ FLE (15)/ PO (6) | TLE (31)/ FLE (9)/ PO (6) | **<0.001** |
| *FBTCS<sup>†</sup> (% of patients)* | 80 (32.4%) | 76 (83.5%) | 59 (35.8%) | **<0.001** | 16 (32.0%) | 23 (88.5%) | 22 (47.8%) | **<0.001** |
| *HS n (% of patients)* | 33 (13.4%) | 13 (14.3%) | 52 (31.5%) | **<0.001** | 9 (18.0%) | 0 | 27 (58.7%) | **<0.001** |
| *Frequent seizures > weekly; n (%)* | 124 (50.2%) | 46 (50.5%) | 125 (75.8%) | **<0.001** | 26 (52%) | 13 (50%) | 42 (91.3%) | **<0.001** |
| *ASM median (range)\** | 2 (0-6) | 2 (0-5) | 2 (0-5) | 0.188 | 2 (1-3) | 2 (1-3) | 2 (1-3) | 0.910 |
| *Surgery n (% of patients) (Localization, n)* | 48 (19.4%)<br>(37 TLE/ 8 FLE/ 3 PLE) | 22 (24.2%)<br>(17 TLE/ 5 FLE) | 38 (23.0%)<br>(31 TLE/ 7 FLE) | 0.538 | 12 (24%)<br>(10 TLE/ 2 PO) | 5 (19.2%)<br>(3 TLE/ 2 PO) | 17 (37.0%)<br>(16 TLE/ 1 PO) | 0.199 |
| **Engel I-II surgical outcome**<br>*n* (% of patients undergone surgery) | 39 (81.3%) | 11 (50%) | 28 (73.7%) | **0.026** | 10 (83.3%) | 2 (40%) | 14 (82.4%) | 0.114 |
| **Post-operative follow-up**<br>*y*, median (*range*) | 7.0 (3-14) | 5.5 (2-15) | 4.8 (3-14) | 0.217 | 5.0 (4-7) | 5.0 (4-5) | 5.0 (3-7) | 0.594 |
| **Sustain Stage**<br>*mean* (*SD*) | 12.5 (14.5) | 8.8 (15.3) | 12.7 (13.7) | 0.072 | 6.7 (6.3) | 7.9 (13.2) | 5.8 (7.1) | 0.650 |
|  | **IGE** |  |  |  |  |  |  |  |
|  | **Discovery cohort** |  |  | **External validation cohort** |  |  |  |  |
|  | **Cortex** | **Basal ganglia** | ***P* value**<br>*(For intergroup differences within cohort)* | **Cortex** | **Basal ganglia** | ***P* value**<br>*(For intergroup differences within cohort)* |  |  |
| **Percentage of total sample** | 78 (40.4%) | 115 (59.6%) | N/A | 42 (68.9%) | 19 (31.1%) | N/A |  |  |
| **Sex** (*F/M*) | 49/29 | 65/50 | 0.600 | 15/27 | 12/7 | <0.001 |  |  |
| **Age**<br>*mean* (*SD*) | 30.8 (8.9) | 35.7 (12.6) | 0.018 | 19.5 (4.4) | 22.5 (7.1) | 0.472 |  |  |
| **Age of onset**<br>*mean* (*SD*) | 13.7 (5.0) | 14.1 (6.8) | 0.840 | 13.3 (2.7) | 14.0 (2.6) | 0.936 |  |  |
| **Duration of epilepsy**<br>*mean* (*SD*) | 22.1 (13.1) | 18.1 (10.5) | 0.059 | 6.1 (4.8) | 8.3 (7.4) | 0.211 |  |  |
| **Syndrome** (*n*) | JAE (27)/ JME (25)/ GTCS-unc (26) | JAE (16)/ JME (21)/ GTCS-unc (78) | <0.001 | JME (42) | JME (19) | N/A |  |  |
| **GTCS†** (% of patients) | 26 (33.3%) | 78 (67.8%) | <0.001 | 7 (16.7%) | 14 (73.7%) | <0.001 |  |  |
| **Frequent seizures**<br>> weekly; *n* (%) | 43 (55.1%) | 52 (45.2%) | 0.400 | 21 (50.0%) | 15 (78.9%) | 0.033 |  |  |
| **ASM**<br>*median* (*range*)* | 2 (0-4) | 2 (0-4) | 0.833 | 1 (0-2) | 1 (0-2) | 0.461 |  |  |
| **Sustain Stage**<br>*mean* (*SD*) | 3.2 (4.2) | 7.0 (8.3) | <0.001 | 4.6 (9.3) | 4.8 (9.8) | 0.962 |  |  |

† Refers to at least one FBTCS (in focal epilepsies)/one GTCS (in IGE) experienced in the year prior to the investigation; \* Kruskal Wallis Test. FBTCS: focal to bilateral tonic-clonic seizure(s); F: female; FLE: frontal lobe epilepsy; HC: healthy control; HS: hippocampal sclerosis; GTCS: generalized tonic-clonic seizure(s); GTCS-unc: IGE unclassified, with GTCS as main seizure subtype; IGE: idiopathic generalized epilepsy; JAE: juvenile absence epilepsy; JME: juvenile myoclonic epilepsy; L: left; M: male; n: number; OLE: occipital lobe epilepsy; PLE: parietal lobe epilepsy; R: right; SD: standard deviation; TLE: temporal lobe epilepsy; U: unknown.

The *hippocampal* subtype mainly included people with TLE (78.2%) and had the highest proportion of mesial TLE with hippocampal sclerosis (TLE-HS; 31.7%) compared to the *cortical* (13.4%) and *basal ganglia* (14.3%) subtypes (χ^2^_2_=20.92, *p*<0.0001; Table 2). Most people included in the *hippocampal* subtype (75.8%) had at least weekly seizures compared to 50.2% and 50.5% in the *cortical* and *basal ganglia* subtypes (χ^2^2=45.99, *p*<0.0001). More people assigned to the *basal ganglia* subtype had focal to bilateral tonic-clonic seizures (FBTCS) in the year before MRI (83.5%, versus 32.4% and 35.8% of *cortical* and *hippocampal* subtypes; χ^2^2=75.93, *p*<0.0001). These findings were virtually identical in the validation cohort (χ^2^2>20.56, all *p*<0.0001). Across progression subtypes, SuStaIn stages were correlated with the duration of epilepsy in the discovery (Spearman’s *ρ*=0.166, *p*<0.0001; Fig. 2f) and external validation cohort (*ρ=*0.268, *p*=0.008; Supplementary Fig. 2), with an increasing weighted stage relating to longer disease duration. Correlations of SuStaIn stages with age at onset (*ρ*=0.036, *p*=0.418) and seizure frequency (*ρ*=-0.013, *p*=0.766) were not statistically significant; the association between SuStaIn stages and the occurrence of FBTCS in the discovery/validation cohorts was also not significant (Kruskal-Wallis *H*= 2.81/1.03, *p*=0.094/0.311). Lastly, in people with a proven lateralised epileptic focus (n=418/122, discovery/validation cohort), ipsilateral cortico-subcortical regions atrophied earlier than contralateral counterparts, irrespective of progression subtype (Supplementary Fig. 3).

We then entered clinical characteristics such as seizure frequency, disease duration, the occurrence of FBTCS, and anti-seizure medications (ASMs) trialled over life into a principal component (PC) analysis, which yielded two PCs with eigenvalues >1 (Supplementary Results): (i) PC1 (32.2% explained variance), with positive loadings of lifetime trialled ASMs and seizure frequency, which we used as a surrogate marker for poorly controlled epilepsy; and (ii) PC2 (26.0% explained variance), with positive loading of epilepsy duration and negative loadings of FBTCS and seizure frequency, which we used as a marker of (chronic) well-controlled epilepsy. Within-individual expression of the hippocampal subtype was associated with less expression of the well-controlled marker (*ρ*=-0.134, *p*=0.003; Fig. 2g); expression of the *cortical* subtype was associated with less expression of the poorly controlled epilepsy marker (*ρ*=-0.320, *p*<0.0001; Fig. 2h), while the opposite relationship held for the expression of the basal *ganglia* subtype (*ρ*=0.204, *p*<0.0001; Fig. 2i).

In the discovery cohort, 21.4 % had epilepsy surgery, and in the validation cohort, 27.9%, with no significant differences among subtypes regarding the proportion of those having surgery (Table 2). In the discovery cohort, people in the *basal ganglia* subtype had lower chances of a good postsurgical seizure outcome (50% Engel classes I-II) compared with those in the *cortical* (81.3%) and *hippocampal* (73.7%) subtypes (χ^2^_2_=7.41, *p*=0.026); within-individual expression of the *basal ganglia* subtype was also negatively correlated with surgical outcome class (Spearman’s *ρ*=-0.238, *p*=0.013). Findings in the validation cohort were qualitatively similar (good outcome: 83.3%, 82.3%, and 40% in *cortical, hippocampal*, and *basal ganglia* subtypes), but group differences (χ^2^_2_=4.337, *p*=0.114) and correlation between within-individual *basal ganglia* subtype expression and surgical outcome class (Spearman’s *ρ*=-0.210, *p*=0.303) were not statistically significant, likely owing to the small surgical sample (n=34).

### SuStaIn identifies two IGE subtypes based on the spatiotemporal progression of atrophy

In IGE, SuStaIn yielded two progression subtypes, with largely overlapping findings in both cohorts (Fig. 3a-c, Fig. 4, Supplementary Fig. 4): a *cortical* subtype, with 40.4% and 68.9% of people in the discovery and validation cohorts (cross-validation folds=0.90, 95% CI=0.82-0.90, discovery cohort, Fig. 3a; 0.79, 95% CI=0.74-0.82, validation cohort); and (2) a *basal ganglia* subtype, including 59.6% and 31.1% of people in the discovery and validation cohorts (cross-validation folds=0.87, 95% CI=0.80-0.88, discovery cohort, Fig. 3b; 0.62, 95% CI=0.60-0.67, validation cohort). There were no differences in subtype distribution between the two cohorts (χ^2^_1_=1.37, *p*=0.504). Spatiotemporal sequences of atrophy in both IGE subtypes were similar to those in focal epilepsy, with temporoparietal regions and the globus pallidus affected first in the *cortical* and *basal ganglia* IGE subtypes.

**Figure 3.**
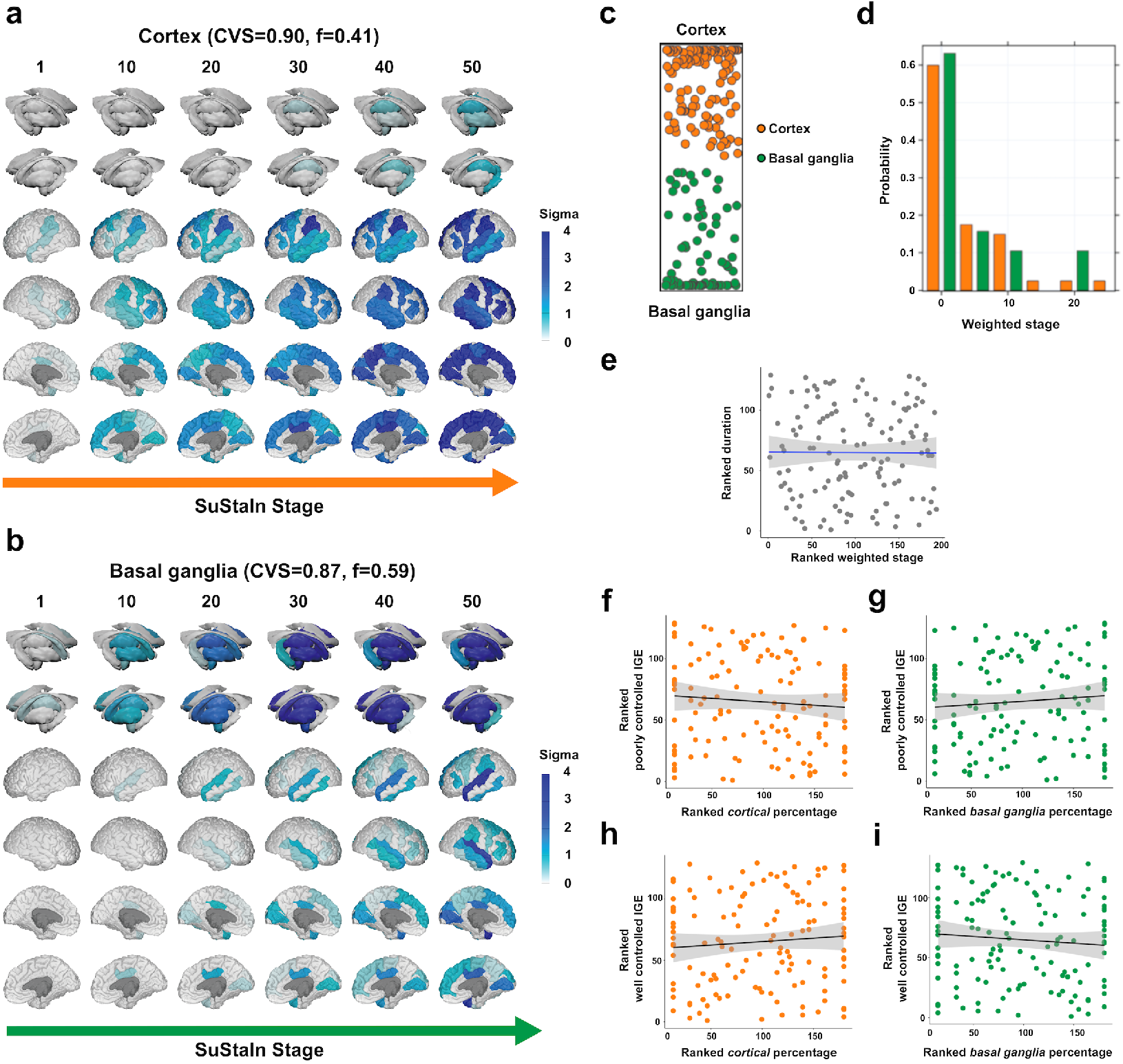
MRI-based progression subtypes in IGE. The figure shows the spatiotemporal patterns of progression of grey matter atrophy (subtypes; **a**. *cortical*; **b**. *basal ganglia*) identified via SuStaIn in the IGE discovery cohort. In panels **a-b**, the colour of each region indicates the severity of grey matter loss; white: unaffected areas; light blue: mildly affected areas (*z*-score=1-2); blue: moderately affected areas (*z*-score=2-3); and dark blue: severely affected areas (*z*-score >3); “CVS’’: cross-validation similarity. “*f*”: proportion of participants assigned to each subtype. Panel **c** shows the assignability of the disease subtype, operationalised as the distance from each vertex of the triangle, whereby each vertex represents the point at which membership of a given subtype is maximal (100%). Panel **d** shows the probability with which each participant from the IGE discovery cohort was assigned a specific SuStaIn stage (stage ranges: 0.005–39.384). Panel **e** shows the correlation between duration of epilepsy and weighted stage, which was not significant. Panels **f-g** show the correlations between within-individual expression of *cortical* and *basal ganglia* subtypes and a marker of poorly controlled IGE (PC1), which were not significant; panels **h-i** show the correlations between within-individual expression of *cortical* and *basal ganglia* subtypes and a marker of well controlled IGE (PC2). Correlation analyses were conducted with Spearman’s *ρ*; the associated panels show ranked data.

**Figure 4.**
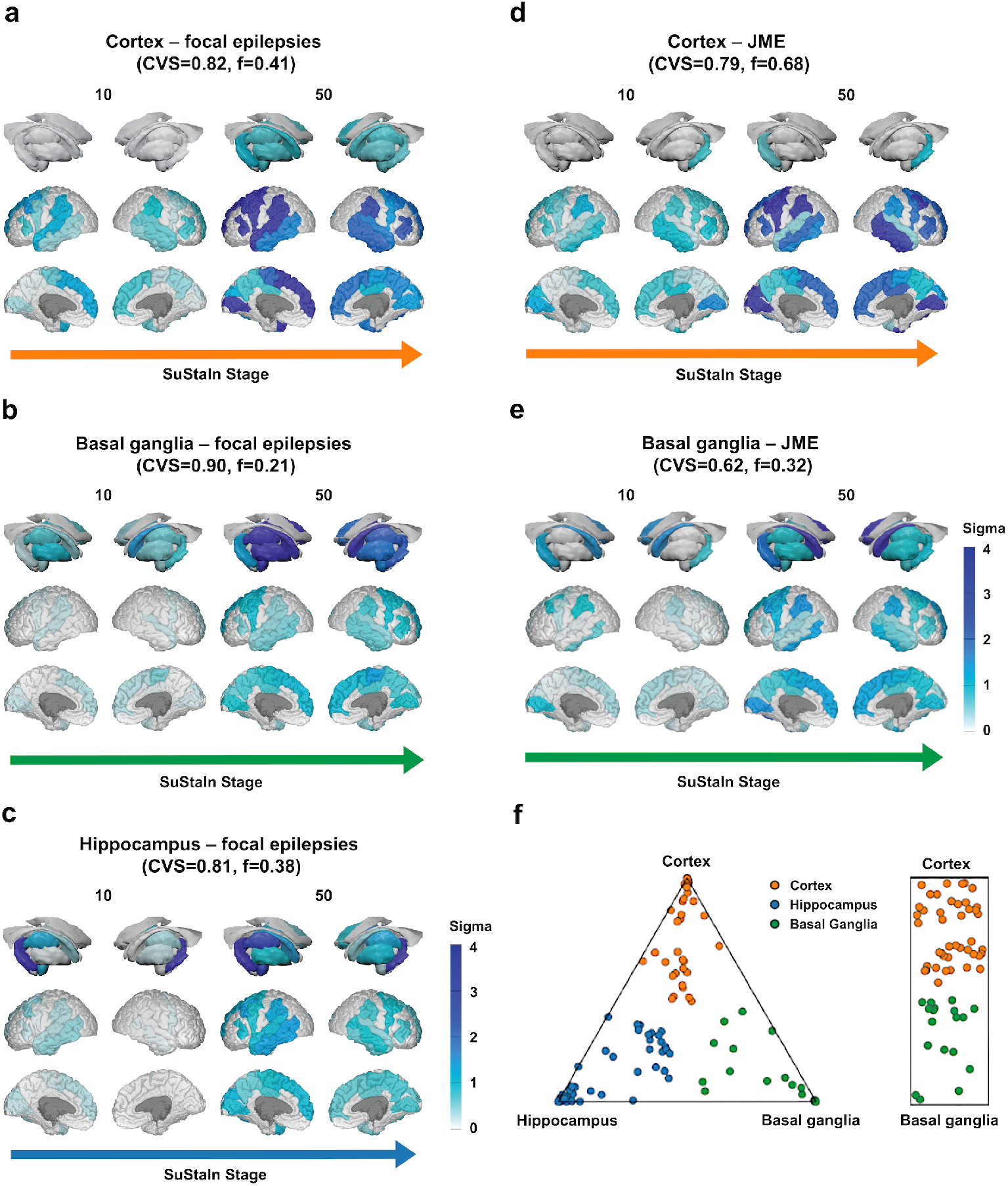
Progression subtypes in the external validation cohorts. The figure shows the atrophy progression subtypes identified via SuStaIn in the focal epilepsy and IGE validation cohorts. Panel **a-c** show the three focal epilepsy subtypes (**a**. *cortical*; **b**. *basal ganglia*; **c**. *hippocampal*), panels **d-e** show the two IGE subtypes (**d**. *cortical*; **e**. *basal ganglia*). In all panels, we show stages 10 and 50 as example stages; the complete progression sequences are provided in Supplementary Fig. 2 (focal epilepsy) and Supplementary Fig. 4 (IGE). The colour of each region indicates the severity of grey matter loss; white: unaffected areas; light blue: mildly affected areas (*z*-score=1-2); blue: moderately affected areas (*z*-score=2-3); and dark blue: severely affected areas (*z*-score >3). Panel **f** shows the probability with which each participant from the focal epilepsy cohort (triangular plot on the left) and IGE cohort (bar plot on the right) was assigned a specific SuStaIn stage (stage ranges: 0.006–54.008, focal epilepsy; 0.004-53.981, IGE). “CVS’’: cross-validation similarity. “*f*”: proportion of participants assigned to each subtype.

### Clinical characterisation of the IGE subtypes

In the IGE discovery group, 66.7% of people assigned to the *cortical* subtype had absence or juvenile myoclonic epilepsy, while 67.8% of those assigned to the *basal ganglia* subtype had unclassified IGE with generalised tonic-clonic seizures (GTCS) as primary seizure type (χ^2^_1_=26.19, *p*<0.0001; Table 2). There were similar findings in the validation cohort: 73.7% of people with JME in the *basal ganglia* subtype had GTCS in the year before MRI, compared to 16.7% of those in the *cortical* subtype (χ^2^_1_=29.06, *p*<0.0001; Table 2). In IGE, SuStaIn stages were not significantly associated with epilepsy duration (*ρ*=0.028/0.08, *p*=0.68/0.18; discovery/validation cohort; Fig. 3e) nor with age of onset (*ρ*=0.036/-0.091, *p*=0.684/0.486, discovery/validation cohort). We then entered clinical characteristics such as GTCS in the year before the scan, epilepsy duration, and ASMs trialled over life into a principal component analysis, that generated two PCs with eigenvalues >1 (Supplementary Material): (i) PC1 (42.0% explained variance), with positive loadings of the number of lifetime trialled ASMs and GTCS in the year before MRI, which we operationalised as a marker of poorly controlled IGE; and (ii) PC2 (34.2% explained variance), with positive and negative loading of the duration of epilepsy and GTCS, which was operationalised as a marker of well-controlled IGE. There were no significant correlations between individual-level *cortex-* or *basal ganglia-subtype* expression and PC1 or PC2 (all *p*>0.35; Fig 3f-i).

## Discussion

We assessed more than 1000 people with epilepsy who underwent high-resolution structural MRI in two specialist epilepsy centres. We used SuStaIn, an established ML algorithm that infers longitudinal sequences of progression of grey matter atrophy from cross-sectional data. Our findings indicate that focal epilepsy and IGE present with latent spatiotemporal patterns of progression, characterised by cortical or basal-ganglia drivers of atrophy, that are differentially co-expressed in each individual. A subtype exclusive to focal epilepsy captured the progression of grey matter damage starting from the hippocampus. Analyses in the external validation cohort corroborated result generalizability. We also identified associations between progression subtypes and markers of disease severity and chronicity, which supports the clinical relevance of our findings. Our study provides dimensional evidence in a categorical framework. It delivers an innovative, imaging- and AI-guided epilepsy taxonomy that may be leveraged for future advancements in individualised prognostics and targeted therapeutics.

Prior studies assessed cumulative grey matter loss in focal epilepsy.^12,13,17^ We show that the reorganization of brain structure over time also affects IGE. Thus, our findings challenge prior views of IGEs as benign disorders,^32^ advocates for future research into strategies to mitigate progression other than surgery. More broadly, our work favours a reconceptualization of the epilepsies as dynamic disorders associated with a deterioration of brain structure, indicating shared features with classical neurodegenerative disorders.^26,28,29,31^ Prior research into cumulative grey matter changes in epilepsy focused on proving its *occurrence* but did not address the temporal signatures of its unfolding. Our work conveys instead a comprehensive overview of spatiotemporal trajectories of progression. It establishes that these (i) imply a complex cortico-subcortical interplay, (ii) are simultaneously co-expressed to a different extent in each individual, and (iii) vary systematically among them. As the set of coordinated changes that underlie progression trajectories is individual-specific, it can thus be viewed as a *spectrum* e.g., as a dimensional entity. Identifying discrete progression subtypes, however, allows us to parse interindividual variability into *categories*, providing a compact classification framework that highlights the main patterns of vulnerability to atrophy. Our application of SuStaIn to uncover progression trajectories links with recent endeavours in other neurological disorders,^26,28,29,31^ and can further inform mechanistic modelsof neurodegeneration. Collectively, we provide strong incentives for accelerated pathways facilitating early diagnostic characterisation along with rapid and effective treatment: *time is brain*.^33^

Progression trajectories involving the cortex or the basal ganglia had broadly similar spatiotemporal characteristics in both focal and generalised epilepsies. Thus, despite the distinct clinical profiles of focal and generalised epilepsies, the substrates of neural vulnerability may be construed as *trans-syndromic*, and prompt a reassessment of the long-standing dichotomisation between focal and generalised epilepsies.^3^ In this context, we echo prior multicentre, cross-sectional evidence for common anatomical signatures,^16,17^ and innovate by proving that these are also dynamic. The overlap in sequences of structural reorganisation may suggest shared pathophysiology. As epilepsies are increasingly conceptualised as network phenomena,^34^ it is plausible that the progression of structural damage may unfold across large-scale, distributed neural networks, as observed in classical neurodegenerative disorders.^35^ Collectively, our work encourages a spectrum-based, *dimensional* conceptualisation of the epilepsies that should complement a purely categorical view, echoing recent endeavours in modern psychiatry.^36,37^

As for the underlying circuitry, atrophy in the *cortical* subtypes first encompasses frontotemporal regions, then progresses fronto-caudally to involve parieto-occipital areas, whose structural alterations were previously shown in cross-sectional work,^17,38^ and affects the hippocampus or subcortical regions only at late stages. The sequence of lateral temporal and frontocentral involvement differed between discovery and validation cohorts in focal epilepsies. This may be explained by differences in the composition of the two cohorts: the discovery cohort had a predominance of people with TLE (∼60%) and a temporo-frontal sequence; in contrast, the validation cohort had people with TLE and FLE in equal proportion and a frontotemporal sequence, which suggests that cortical areas near the epileptogenic networks may be affected first. In support of this view, we also observed that people with focal epilepsy had an earlier involvement of ipsilateral regions. Our interpretation reiterates our previous findings in a longitudinal study^13^ and evidence of graded severity of diffusion abnormalities in TLE as a function of the Euclidean distance from the seizure focus.^39^ Future work may benefit from larger samples of people with extratemporal epilepsies and people with well-established seizure focus lateralisation and localisation to disentangle their influence on progression. In IGE, progression trajectories in the *cortical* subtype broadly recapitulated those of focal epilepsies, with minor differences between the discovery and validation cohorts that may stem from heterogeneity in diagnostic characteristics. Half of the people in the discovery cohort had a diagnosis of unclassified IGE, while people in the validation cohort had JME. Our findings in IGE implicate areas previously identified as atypical in cross-sectional imaging studies.^40–43^ Longitudinal studies investigating trajectories of structural reorganisation in IGE are rare. In JME, altered development of association cortices occurs in the first two years after diagnosis, with an attenuated age-related decline in thickness and surface area compared to typical neurodevelopment.^44^ Such evidence was obtained in a paediatric cohort, while our IGE samples had an average age of >20 years. Future research is thus needed to characterise lifelong patterns of cortico-subcortical reorganisation in IGE and compare trajectories in childhood and adolescence to those in adulthood.

We identified a *basal ganglia* progression subtype in focal and generalised epilepsies, which showed similar patterns of spatiotemporal evolution, first involving the globus pallidus, then the caudate and thalamus, followed by cortical areas. People with focal epilepsy and predominance of this subtype were more likely to have FBTCS, while those with IGE were more likely to be unclassified IGE with GTCS. Multimodal evidence from animal models and humans implicates thalamocortical and basal ganglia circuitry in the generation or modulation of tonic-clonic seizures.^15,45–50^ In IGE, imaging studies have documented subcortical grey matter volume loss, particularly in the thalamus,^15,51^ and reorganisation of structural and functional thalamocortical connectivity.^18^ EEG-fMRI studies also implicated the thalamus in the generating generalised spike-wave discharges and absence of seizures, and showed hyperconnectivity among basal ganglia.^52^ In TLE, multimodal evidence points to thalamic atrophy and reorganization of thalamic and basal ganglia connectivity,^16,17,53–55^ which appears more marked in TLE with FBTCS.^46,47,56,57^ Collectively, our findings link with prior evidence on the role of the thalamus and basal ganglia in tonic-clonic seizures, pointing to substantial similarities in the pathophysiology of focal and generalized epilepsy. We postulate that the circuitry involved in tonic-clonic ictogenesis undergoes structural damage more precociously.

In focal epilepsies, we also identified a progression subtype characterised by initial involvement of the hippocampus, followed by the thalamus and temporal neocortex, and subsequently by other cortical areas. The spatial distribution of this progression pattern resembles that of areas exhibiting grey matter alterations and implicated in ictogenesis in TLE, particularly TLE-HS.^16,17^ Our findings mirror the results of single-centre longitudinal imaging studies^13,58,59^ and our meta-analysis,^12^ showing progressive thalamic and hippocampal atrophy and demonstrated that areas affected by the progression of atrophy^13^ were structurally connected to the hippocampus. We thus reiterate that seizure onset, propagation and progressive brain damage may be closely linked, with regions preferentially implicated in seizures deteriorating first. Notably, not all people with the predominant *hippocampal* progression subtype had TLE. The hippocampus may exhibit heightened vulnerability to developmental, hormonal and environmental factors, which is corroborated by its involvement in a broad spectrum of brain disorders.^60–62^ Studies in healthy adults identified the hippocampus as a component of a late-developing brain network with significant vulnerability to the effects of ageing and disease,^35^ and prior investigations of extratemporal epilepsy documented subtle structural hippocampal alterations such as hippocampal malrotation.^63,64^ It is possible that repeated seizures, even if not mesiotemporal, may ultimately lead to pathological mesiotemporal reorganisation, which could then propagate to other regions following the main axes of hippocampal structural connectivity.

Correlation analyses contextualised the identified epilepsy progression subtypes from a clinical viewpoint. In focal epilepsies, the duration of disease, but not age at onset, was associated with subtype staging. While replicating prior cross-sectional evidence,^12,17^ our findings indicate that the progression of grey matter damage along the topographical axes captured by each subtype is time-dependent but may not be substantially influenced by the developmental stage at diagnosis. Seizure frequency was higher in those with predominant *hippocampal* progression. It is tempting to speculate that a higher seizure burden may more severely affect the hippocampus and lead to a progression cascade involving interconnected areas, per our prior considerations on hippocampal and network-level vulnerability to disease. Correlation analyses cannot establish causal chains, which need validation in future longitudinal studies. Principal component analyses showed associations between epilepsy severity and progression subtypes, suggesting more aggressive disease in the cortical and basal ganglia subtypes. These findings show how SuStaIn may be used for clinical stratification, with pertinent implications. As subtype expression and their combination are quantifiable within individuals, findings of higher cortical and basal ganglia loading have prognostic implications and may prompt accelerated treatment pathways. Similarly, people with focal epilepsy and predominant basal ganglia-led progression benefited less from epilepsy surgery, which may stem from a higher burden of secondary generalisation, an established predictor of unfavourable post-surgical outcome;^65–67^ the latter finding can also be translated to clinical decision making. In IGE, people with uncontrolled GTCS were preferentially assigned to the *basal ganglia* subtype. Still, we did not otherwise identify significant correlations between clinical characteristics and subtypes and their stages. The underlying determinants of progression in IGE and focal epilepsies may differ, despite the considerable overlap in neuroanatomical signatures. IGE is characterised by a complex polygenic aetiology,^68,69^ which may be an essential driver of interindividual differences in the expression of progression subtypes and their associations with clinical phenotypes. These hypotheses will require validation in future imaging-genetics investigations.

A strength of our work is the inclusion of an external validation cohort, which strongly supports generalizability. SuStaIn is an open-source algorithm widely applied to multicentre cohorts of people with neurodegenerative disorders, it only requires cross-sectional datasets to detect multiple spatiotemporal trajectories and provides probabilistic and quantitative data information for individualised inference.^26–28,31^ Thus, we employed state-of-the-art, previously validated methods to maximise reproducibility and replicability. Cortical thickness, hippocampal and subcortical volumes can be reliably and non-invasively quantified using structural MRI and are validated morphometric markers of neuronal loss.^16,17^ One limitation is the use of a parcellation scheme that does not cover the whole brain. However, we note that the selection of regions used here was attained to maximise the trade-off between accuracy and computational complexity and was motivated by the findings of large-scale multicentre studies of the ENIGMA-Epilepsy consortium.^16,17^

In conclusion, we evaluated over a thousand people with epilepsies using an unsupervised ML algorithm and routinely acquired structural MRI scans. We provide a novel epilepsy taxonomy based on the differential spatiotemporal progression of grey matter atrophy. Progression subtypes principally implicate neocortical and basal ganglia drivers both in focal and generalised epilepsies, and limbic circuitry in focal epilepsy only. They are differentially co-expressed in each individual, and relate to clinical indicators of disease severity. Classification of people with epilepsy capitalises on the maximally expressed progression subtype at the personal level, conveys a dimensional perspective into a categorical framework, and conceptually advances the extant categorical classification approaches. By providing an individual-level characterisation of the underlying biology, we offer deliverables that can be used prospectively to enhance individualized prognostic and therapeutic considerations and may aid clinical stratification for future clinical trials of disease-modifying agents.

## Methods

### Participants

We analysed anonymised data from two separate cohorts. The discovery cohort consisted of structural MRI data of a single-centre dataset involving long-term follow-up^70^ of individuals with focal epilepsy or IGE, investigated with 3T high-resolution MRI (GE, Milwaukee, WI, USA) at the Chalfont Centre for Epilepsy (UCL Queen Square Institute of Neurology/National Hospital of Neurology and Neurosurgery, London), UK, between January 2004 and March 2018. An external, independent validation cohort included individuals scanned with 3T high-resolution MRI at the Department of Neurology of West China Hospital, Sichuan University, Chengdu, China, between June 2013 and December 2020. After visual inspection detailed in supplementary material, 894/211 people with epilepsy and 121/73 control participants, for a total of 1299 scans, were considered for inclusion in this work.

Diagnosis, lateralisation, and localisation of focal epilepsy were made by a multidisciplinary epilepsy team, based on clinical history, neurological examination, seizure semiology, ambulatory EEG monitoring, interictal and ictal EEG during long-term video-EEG telemetry, structural MRI, [18F]fluorodeoxyglucose PET (in a subset), and neuropsychological assessments. We excluded people with brain lesions other than hippocampal sclerosis, those with poor data quality, and those without adequately detailed clinical data. People with IGE had a typical clinical history and a previous routine EEG with interictal generalised (poly-)spike-and-wave discharges at UCL. All participants had a clinical structural MRI scan as part of diagnostic investigations. Clinical characteristics were collected through a review of the entry in medical notes closest in time to the date of the MRI scan. The number of lifetime trialled ASMs and the duration of epilepsy were recorded on the day of the MRI scan. Complete details are provided in Table 1. Controls from both sites were recruited from the local community and had no family history of epilepsy or neurological or psychiatric disorders.

### Standard protocol approvals, registrations, and participant consent

This study was pursued under a protocol approved by the UCL and University College London Hospital Joint Research Ethics Committee (20/LO/0149) and involved the analysis of previously acquired clinical data posing no risk to patients and did not require individual patient consent. The recruitment of healthy controls was approved by the UCL and University College London Hospital Joint Research Ethics Committee as part of previous studies. Written informed consent was obtained from healthy control participants per the Declaration of Helsinki standards. Participant recruitment in relation to validation cohort data was approved by the West China Hospital Clinical Trials and Biomedical Ethics Committee of Sichuan University. All participants from Sichuan University gave written informed consent by the standards of the Declaration of Helsinki.

### MRI data acquisition

For UCL participants scanned between January 2004 and March 2013 (all people with IGE and 48 controls), MRI data were acquired on a 3T GE Signa HDx scanner with a coronal T1-weighted 3D inversion recovery fast spoiled gradient echo (IR-FSPGR) sequence, repetition time (TR)/echo time (TE)/inversion time (TI): 8.1/3.1/450 ms, voxel size: 0.9×0.9×1.1mm. For UCL participants scanned between March 2013 and March 2018 (all people with focal epilepsy and 70 healthy controls), MRI data were acquired on a 3T GE Discovery MR750 scanner using a 3D T1-weighted magnetisation prepared rapid acquisition gradient echo (MPRAGE) sequence with TE/TR/TI: 3.1/7.4/400 ms, voxel size: 1.0×1.0×1.0 mm. For participants in the external validation cohort, MRI data were acquired at the West China Hospital of Sichuan University between June 2013 and December 2020, using a 3T Siemens Tim Trio MRI scanner (Erlangen, Germany) with an eight-channel head coil. High-resolution T1-weighted MRI was acquired using a 3D MPRAGE sequence with TR/TE/TI: 1900/2.6/900 ms, voxel size: 1.0×1.0×1.0 mm.

### MRI data preprocessing

To evaluate brain atrophy, we focused on cortical thickness as an established, surface-based marker of cortical morphology, that reflects cellular-level features including size, density, and arrangement of neurons, glia, and nerve fibres.^71,72^ To this end, we employed the Computational Anatomy Toolbox (CAT12) running in SPM12 and Matlab 2021a (Mathworks).^73,74^ Cortical thickness was estimated using the projection-based thickness method, previously validated using spherical and brain phantoms confirming accurate measurements under a comprehensive set of parameters for several thickness levels.^73^ The CAT12 toolbox provided excellent test-retest reliability (*R*^*2*^=0.986) and was validated against other cortical surface reconstruction methods, showing fewer measurement errors than similar software.^74,75^ We used an inverse-consistent longitudinal surface registration approach implemented in CAT12 to prevent an asymmetry bias from arising when data from multiple time points are analysed. All data were quality controlled according to standardised CAT12 pipelines; scans that exhibited misalignment, misregistration, or inaccurate thickness estimation were excluded. Image quality ratings were estimated by scaling image noise, inhomogeneities, and resolution to a single score within the CAT12 quality assurance framework. Following quality checks, 80/28 participants with epilepsy and 1/2 controls in the discovery/validation cohorts were excluded. Detailed demographic and clinical characteristics of individuals retained for SuStaIn analysis are provided in Table 1.

To obtain measures of hippocampal volume, we employed Hipposeg (http://niftyweb.cs.ucl.ac.uk/program.php?p=HIPPOSEG), an open-source, multi-atlas-based, previously-validated hippocampal segmentation algorithm.^76^ Hipposeg delineates the hippocampus with no more variability than seen between expert human raters, and is robust to hippocampal morphological alterations, including atrophy.^77^ Volumes of other subcortical structures relevant in epilepsy, including the thalamus, amygdala, caudate, putamen, and globus pallidus, and total intracranial volume (TIV), were extracted using a parcellation algorithm based on Geodesic Information Flows (GIF),^78^ freely available within NiftyWeb (http://cmictig.cs.ucl.ac.uk/niftyweb, UCL Centre for Medical Image Computing, UK). Adjustment for TIV is described below. Previous work showed excellent agreement between GIF-derived subcortical volumes and those obtained using FSL-FIRST,^79^ and between GIF-derived cortical volumes and those obtained using SPM12.^80^

### Regions of interest specification

Based on the recent international multicentre ENIGMA-epilepsy structural MRI study,^17^ we selected the following bilateral regions of interest (ROI) of the Desikan-Killiany (DK40) atlas: (i) 28 cortical ROIs: left and right superior frontal gyrus, caudal middle frontal gyrus, inferior frontal gyrus– pars *triangularis*, precentral gyrus, paracentral lobule, superior temporal gyrus, transverse temporal gyrus, middle temporal gyrus, inferior temporal gyrus, supramarginal gyrus, precuneus, posterior cingulate cortex, lingual gyrus, and cuneus; and (ii) 12 mesiotemporal and subcortical ROIs, including left and right hippocampus, amygdala, thalamus, and basal ganglia structures, including caudate, globus pallidus, and putamen (Fig. 1a; Supplementary Figure 1). While the above parcellation scheme was constrained to a maximum of 40 regions of interest and did not allow for whole-brain inference, as in prior work with SuStaIn,^26,30,31^ we note that the ROI selection used in our study was attained to maximise the trade-off between accuracy and computational tractability, and was motivated *a priori* by the findings of large-scale multicentre studies of the ENIGMA-epilepsy consortium,^16,17^ which provided a state-of-the-art characterisation of the spatial distribution of grey matter alterations in focal epilepsy and IGE.

As in prior structural MRI investigations employing the SuStaIn algorithm,^26,30,31^ we adjusted ROI-wise cortical thickness within each cortical ROI for age at the scan and (binary) sex and adjusted mesiotemporal and subcortical volumes for total intracranial volume, age at scan, and sex; specifically, we constructed a linear regression model for each region separately, entering the value of cortical thickness and subcortical volumes as the dependent variable and the variables mentioned above as predictors, and retained the unstandardised residuals (of the fit) for each region for subsequent analyses. For each of the 40 MRI measures listed above, we combined the two healthy control datasets to fit a Bayesian linear regression model with total intracranial volume, sex, age and age squared as independent variables, and each MRI measure as the outcome.^26,30,31^ As previously,^26,30,31^ we computed the expected values using this model and subtracted the observed values to obtain residual values of each MRI variable. We refer to the residual values as adjusted values.^26,30,31^ To investigate the effects of seizure focus laterality in people with focal epilepsies, we conducted a subgroup analysis by including only people with proven lateralisation of the seizure focus and regrouping regions into “ipsilateral” or “contralateral”.

### Subtype and Stage Inference (SuStaIn)

As in previous work,^26,28,31^ we employed the Subtype and Stage Inference algorithm (SuStaIn)^26^ to identify distinct patterns of spatiotemporal progression from cross-sectional imaging data, coded as a set of stages, that are co-expressed to a different extent in each individual.^19^ SuStaIn simultaneously clusters individuals into groups (*subtypes*), based on the predominant expression of a given progression pattern. Detailed formalisation and mathematical modelling of SuStaIn has been published previously^26,28^ and will not be extensively detailed here. In Fig. 1, we provide a conceptual overview of the application of the SuStaIn algorithm in our study and dedicate the following paragraphs to briefly overview the methodology and outline parameter choices specific to our analyses. Notably, we used the SuStaIn algorithm separately for focal epilepsies and IGE. We also repeated the same analyses in the external validation cohort data.

We used SuStaIn with the *piecewise linear z-score model* of disease progression to estimate the most likely sequence with which selected regions reach different atrophy levels over time (Fig. 1b), *i*.*e*., to identify spatiotemporal atrophy patterns. Each progression pattern is described as a series of *stages*, whereby each stage corresponds to a biomarker (cortical thickness or volume of a brain region) reaching a new *z*-score. The optimal number of subtypes was determined using information criteria calculated through cross-validation^81^ to balance model complexity with internal model accuracy, as described previously.^26^ Briefly, the piecewise linear *z*-score model requires *z*-scored data as input. Thus, each regional volume measurement was expressed as a *z*-score relative to the control group by normalising each dataset relative to its control population in each institution, so that the control population had a mean of 0 and standard deviation of 1.^26^ As the UCL cohort consisted of participants acquired with two different scanners (3T GE Signa, “old scanner”; and GE Discovery, “new scanner”; details above), people imaged with the old/new scanner were normalised to controls imaged with the old/new scanner only. In the context of progressive, disease-associated atrophy, regional cortical thinning and subcortical volumes decrease over time; thus, regional *z*-scores also become negative as a disease progresses. The piecewise linear *z*-score model, however, requires that *z*-scores *increase* as a function of disease progression. Hence, we multiplied the above-obtained *z*-score by -1 to allow for model fit, as previously.^26^ We then ran the SuStaIn algorithm with 25 start points and 1,000,000 Markov Chain Monte Carlo (MCMC) iterations, as previously,^26^ and evaluated solutions up to a maximum of *n*=4 clusters (Fig. 1b); the data-driven output of the SuStaIn algorithm, run separately in focal epilepsies and IGE, subsequently indicated inability to identify clusters beyond *n*>3 (focal epilepsies) and *n*>2 (IGE). We then performed 10-fold cross-validation (Fig. 1b) to evaluate the optimal number of clusters that best describes unseen data and assess the stability of subtype progression patterns across folds; the cross-validation similarity metric for the progression subtypes across validation folds ranges from 0 (no similarity) to 1 (maximum similarity).^26^ SuStaIn calculates the probability with which each individual may fall into each stage of each subtype, with subtype expression ranging from 0 to 1; individuals are then assigned to their maximum likelihood subtype and stage. Individuals with no abnormalities in thickness or volume in any region are classified by SuStaIn as “weighted stage 0” and are not assigned to a progression subtype. In our study, there were no individuals assigned to weighted stage 0. We then quantified the proportion of individuals classified into each subtype. We staged individuals by computing their average SuStaIn stage, weighted by the probability that they belonged to each stage of each subtype.^26^

### Statistical analysis

Data were analysed using IBM SPSS version 26 and R 4.2.1. For analysis of demographic and clinical data, we used ANOVA, Kruskal-Wallis and chi-squared tests for continuous parametric, non-parametric and categorical data, respectively. We conducted a principal component analysis to investigate associations between progression patterns in focal and generalised epilepsies and clinical characteristics. In detail, FBTCS occurring in the year before MRI,^47,56^ seizure frequency, ASMs trialled over life, and epilepsy duration were entered in a PCA for people with focal epilepsy. In contrast, GTCS in the year before MRI, seizure frequency, ASMs trialled over life, and epilepsy duration were entered in a PCA in IGE. We then probed associations of progression patterns with the extracted principal components, which represent superordinate markers of disease severity, using two-tailed nonparametric correlation analyses.

## Data Availability

Anonymized statistical data to reproduce the main findings are available from the corresponding author upon reasonable request from any qualified investigator. Other data are not available due to their containing information that could compromise the privacy of research participants. 

## Acknowledgements

We thank the people with epilepsy and the controls for participation, and Drs Christian Vollmar and Maria Centeno for recruiting part of the controls. This work was supported by the Epilepsy Society, Chalfont St Peter, UK and was carried out at the National Institute for Health Research University College London Hospitals Biomedical Research Centre, which receives a proportion of funding from the UK Department of Health’s Research Centres funding scheme. FX is supported by a Newton International Fellowship of the Academy of Medical Sciences and the Newton Fund (NIF\R5\264) and acknowledges support from the National Natural Science Foundation of China (82001369). LC acknowledges support from Brain Research UK (award n. 14181). BW acknowledges support from the MRC (MR/T005335/1). ALY is supported by an MRC Skills Development Fellowship (MR/T027800/1). Part of participant recruitment was funded through Wellcome Trust awards (project grants 083148 and 079474) to JD and MK. JD acknowledges support from the NIHR and Wellcome Trust (218380). GPW was funded by the MRC (G0802012, MR/M00841X/1). DCA acknowledges support by the Wellcome Trust (221915) for work in this area. BK is supported by the National Institute for Health Research Biomedical Research Centre at UCLH and UCL. JWS receives support from the Dr Marvin Weil Epilepsy Research Fund and the Christelijke Verenigingvoor de Verpleging van Lijdersaan Epilepsie, Netherlands. The authors also acknowledge the facilities and scientific and technical assistance of the National Imaging Facility, a National Collaborative Research Infrastructure Strategy (NCRIS) capability, at the Centre for Microscopy, Characterisation, and Analysis, the University of Western Australia.

## Data availability statement

Anonymised statistical data to reproduce the main findings are available from the corresponding author upon reasonable request from any qualified investigator. Other data are not available due to their containing information that could compromise the privacy of research participants.

## Code availability statement

Python and MATLAB implementations of the SuStaIn algorithm are available on the UCL-POND GitHub page: https://github.com/ucl-pond.

## Citation Diversity Statement

Recent work in several fields of science has identified a bias in citation practices such that papers from women and other minority scholars are under-cited relative to the number of such papers in the field (doi: https://doi.org/10.1016/j.neuron.2020.05.011; https://github.com/dalejn/cleanBib). Here we sought to proactively consider choosing references that reflect the diversity of the field in thought, form of contribution, gender, race, ethnicity, and other factors. First, we obtained the predicted gender of the first and last author of each reference by using databases that store the probability of a first name being carried by a woman. By this measure (and excluding self-citations to the first and last authors of our current paper), our references contain 12.26% woman(first)/woman(last), 22.49% man/woman, 22.49% woman/man, and 42.77% man/man. This method is limited in that a) names, pronouns, and social media profiles used to construct the databases may not, in every case, be indicative of gender identity and b) it cannot account for intersex, non-binary, or transgender people. Second, we obtained predicted racial/ethnic category of the first and last author of each reference by databases that store the probability of a first and last name being carried by an author of color. By this measure (and excluding self-citations), our references contain 12.64% author of color (first)/author of color(last), 18.89% white author/author of color, 21.24% author of color/white author, and 47.22% white author/white author. This method is limited in that a) names and Florida Voter Data to make the predictions may not be indicative of racial/ethnic identity, and b) it cannot account for Indigenous and mixed-race authors, or those who may face differential biases due to the ambiguous racialization or ethnicization of their names. We look forward to future work that could help us to better understand how to support equitable practices in science.

## Notes

**Conflicts of interest:** JWS reports personal fees as a speaker or consultant from Arvelle, Eisai, GW Pharmaceuticals, UCB Pharma, and Zogenix. The other authors report no conflicts of interest in relation to this submission.

### Competing Interest Statement

JWS reports personal fees as a speaker or consultant from Arvelle, Eisai, GW Pharmaceuticals, UCB Pharma, and Zogenix. The other authors report no conflicting interests in relation to this submission.

