## Supplementary Material for "Artificial intelligence and MRI: the source of a new epilepsy taxonomy"

### Supplementary Methods

#### *MRI data: quality control and participant exclusion*

For MRI data of the discovery cohort, acquired at UCL, scan quality was guaranteed by UCL radiographers in terms of whole-brain coverage, absence of artefact and signal losses; the scans were inspected and clinically reported by multiple experienced neuroradiologists. Scans which were labelled as poor in quality either in the radiographers' notes or by the neuroradiologists were not considered for this study. For MRI data of the validation cohort, authors DA, WYL, and DZ carried out quality control checks for the scans of people with focal epilepsy, while DA and YZ carried out those for the scans of people with JME. All individuals with scans of insufficient quality were not considered for this study. Before MRI data preprocessing, a total of 894 people with epilepsy [691 with focal epilepsy (336/355 old/new UCL scanner) and 203 IGE (all old scanner)] and 121 healthy control participants (50/71 acquired with the old/new UCL scanner) from the discovery cohort, and 211 people with epilepsy (150 with focal epilepsy/61 with IGE) and 73 control participants from the validation cohort, for a total of 1299 scans, were considered for inclusion in this work.

The Computational Anatomy Toolbox (CAT) introduces a quality control framework and allows evaluating essential image parameters such as noise, signal inhomogeneities, and image resolution. All these quality measures are subsequently used in the context of a rating scale system that enables the comparison of measures across different scanners and sequences. Moreover, quality measures are summarised into a single quality rating with the following possible options: excellent, good, satisfactory, sufficient, critical, failed. Only scans rated "excellent" or "good" were considered for inclusion and visually inspected, while those not reaching these quality thresholds were discarded. In the discovery cohort, 540 scans of people with focal epilepsy and 195 with IGE were rated as "excellent" or "good", while 151 scans of people with focal epilepsy and 10 scans of people with IGE (all acquired with the old UCL scanner) had inferior quality and were excluded; moreover, an additional 37 scans of people with focal epilepsy, that were rated as "good" but were quantitatively on the low side of the CAT12 "good" rating spectrum, were discarded after visual inspection. The scans of 3 controls (2 old scanner/1 new scanner) were also excluded for low quality ratings. Thus, the final participant number of the discovery cohort consisted of 503 people with focal epilepsy, 193 with IGE, and 118 healthy control participants. In the validation cohort, 122 scans of people with focal epilepsy, 61 (all) scans of people with IGE and 71 of 73 control scans were rated as "excellent" or "good", while 28 scans of people with focal epilepsy and 2 control scans were excluded for lower quality ratings. Thus, the final participant number of the validation cohort consisted of 122 people with focal epilepsy, 61 people with IGE, and 71 healthy controls.

### Supplementary Results

#### *Principal Component Analysis*

These analyses were conducted on discovery cohort data. To further assess the clinical relevance of MRI-based progression subtypes, we first derived composite clinical constructs by entering clinical characteristics into a principal component analysis (PCA), and then assessed the relationship between the former and within-individual subtype expression using Spearman's rank correlations. In focal epilepsy, a PCA based on seizure frequency, disease duration, the occurrence of FBTCS (entered as binary variable: yes/no), and antiseizure medications (ASMs) trialled over life yielded two principal components (PCs) with eigenvalues >1: (i) PC1 (eigenvalue=1.29, 32.2% of explained total variance), with positive loadings of lifetime trialled ASM (0.826) and seizure frequency (0.584), which we operationalized as a superordinate marker of poorly controlled (i.e., chronic and active) epilepsy; and (ii) PC2 (eigenvalue=1.04, 26.0% of explained total variance), with positive loading of epilepsy duration (0.797) and negative loading of FBTCS (-0.613) and seizure frequency (-0.169), which we operationalized as a superordinate marker of (chronic) well controlled epilepsy. In IGE, a PCA on disease duration, occurrence of GTCS in the year before MRI, ASMs trialled over life yielded two PCs with eigenvalues >1: (i) PC1 (eigenvalue=1.19, 42.0% of explained total variance), with positive loading of number of lifetime trialled ASMs (0.645) and GTCS in the year before MRI (0.505), which we operationalized as a superordinate marker of poorly controlled IGE; (ii) and PC2 (eigenvalue=1.03, 34.2% of explained total variance), with a positive loading of duration of epilepsy (0.798) and negative loading of GTCS (-0.58), which we operationalized as a marker of chronic well controlled IGE. Statistical details regarding correlation analyses are provided in the main manuscript text; the associated correlation scatterplots are displayed in Fig. 2 and Fig. 3.

### Supplementary Table 1. Regions of Interest.

| ***ROI types*** | ***Brain areas*** | ***Preprocessing*** |
|---|---|---|
| Cortical thickness (Desikan-Killiany Atlas, DK40) | Left caudal middle frontal | Regional cortical thickness values were computed after correction for age and sex and were transformed to *z*-scored relative to a control population. For the UCL discovery cohort, patient data acquired via the old/new scanner was *z*-scored based on controls acquired with the old/new scanner, respectively. |
| | Right caudal middle frontal | |
| | Left cuneus | |
| | Right cuneus | |
| | Left inferior temporal | |
| | Right inferior temporal | |
| | Left lingual | |
| | Right lingual | |
| | Left middle temporal | |
| | Right middle temporal | |
| | Left paracentral | |
| | Right paracentral | |
| | Left pars triangularis | |
| | Right pars triangularis | |
| | Left precentral | |
| | Right precentral | |
| | Left precuneus | |
| | Right precuneus | |
| | Left posterior cingulate | |
| | Right posterior cingulate | |
| | Left superior frontal | |
| | Right superior frontal | |
| | Left superior temporal | |
| | Right superior temporal | |
| | Left supramarginal | |
| | Right supramarginal | |
| | Left transverse temporal | |
| | Right transverse temporal | |
| Hippocampal volume (Hipposeg) and subcortical volumes (Geodesic Information Flow) | Left hippocampus | Regional volumetric values were computed after correction for age, sex, and total intracranial volume (TIV; head size correction purposes) and were transformed to *z*-scored relative to a control population. For the UCL discovery cohort, patient data acquired via the old/new scanner was *z*-scored based on controls acquired with the old/new scanner, respectively. |
| | Right hippocampus | |
| | Left amygdala | |
| | Right amygdala | |
| | Left caudate | |
| | Right caudate | |
| | Left pallidum | |
| | Right pallidum | |
| | Left putamen | |
| | Right putamen | |
| | Left thalamus | |
| | Right thalamus | |

**Supplementary Figure 1. Cortical and subcortical regions of interest.**

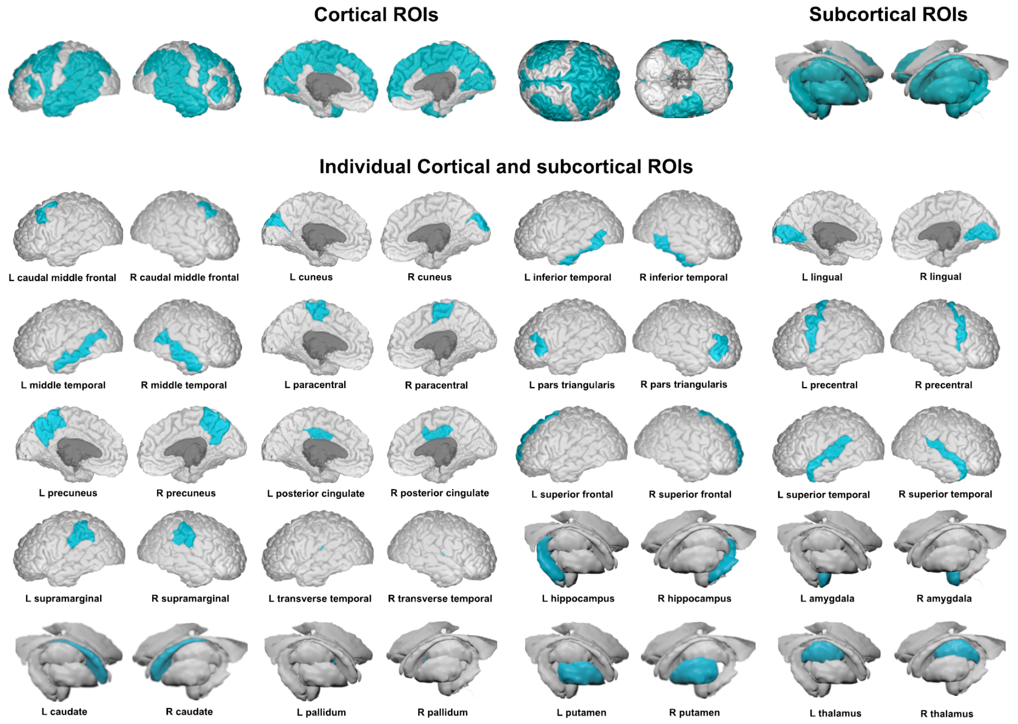


We selected 20 bilateral regions of interest (ROI) of the Desikan-Killiany (DK40) atlas: (a) 14 bilateral cortical regions, including superior frontal gyrus, caudal middle frontal gyrus, inferior frontal gyrus– pars *triangularis*, precentral gyrus, paracentral lobule, superior temporal gyrus, transverse temporal gyrus, middle temporal gyrus, inferior temporal gyrus, supramarginal gyrus, precuneus, posterior cingulate cortex, lingual gyrus, and cuneus; and (b) bilateral ROIs for hippocampus, amygdala, thalamus, and basal ganglia structures, including caudate, globus pallidus, and putamen. The above ROI selection was largely based on the findings of the recent international multicentre ENIGMA-epilepsy structural MRI study.[17] Abbreviations: L= left; R= right; ROI= region of interest.

**Supplementary Figure 2. Progression subtypes in the focal epilepsy external validation cohort.**

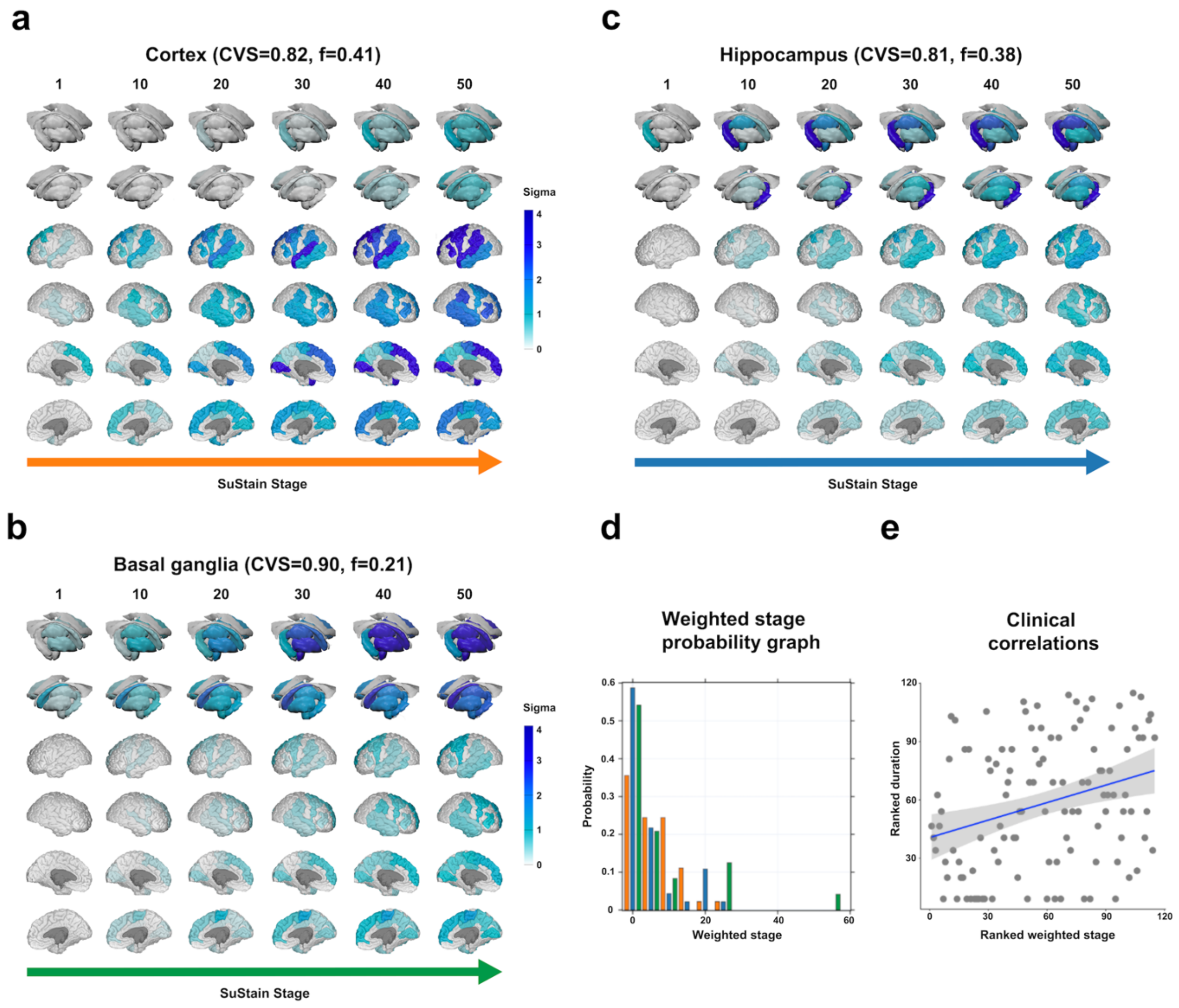


The figure shows the spatiotemporal patterns of progression of grey matter atrophy (subtypes; **a.** *cortical*; **b.** *basal ganglia*; **c.** *hippocampal*) identified via SuStaIn in the focal epilepsy validation cohort. Each progression pattern in panels **a-c** consists of a sequence of stages with which cortical thickness and subcortical volumes reach different z-scores in patients relative to healthy controls. The colour of each region indicates the severity of grey matter loss; white: unaffected areas; light blue: mildly affected areas (*z*-score=1-2); blue: moderately affected areas (*z*-score=2-3); and dark blue: severely affected areas (*z*-score >3); "CVS": cross-validation similarity. "*f*": proportion of participants assigned to each subtype. Panel **d** shows the probability with which each participant from the focal epilepsy discovery cohort was assigned a specific SuStaIn stage (stage ranges: 0.006–54.008). Panel **e** shows the correlation between duration of epilepsy and weighted stage (Spearman's $\rho$=0.268, $p$=0.008), with an increasing weighted stage relating to longer disease duration; the associated panels show ranked data

**Supplementary Figure 3. Progression subtypes in people with focal epilepsy and unilateral seizure focus.**

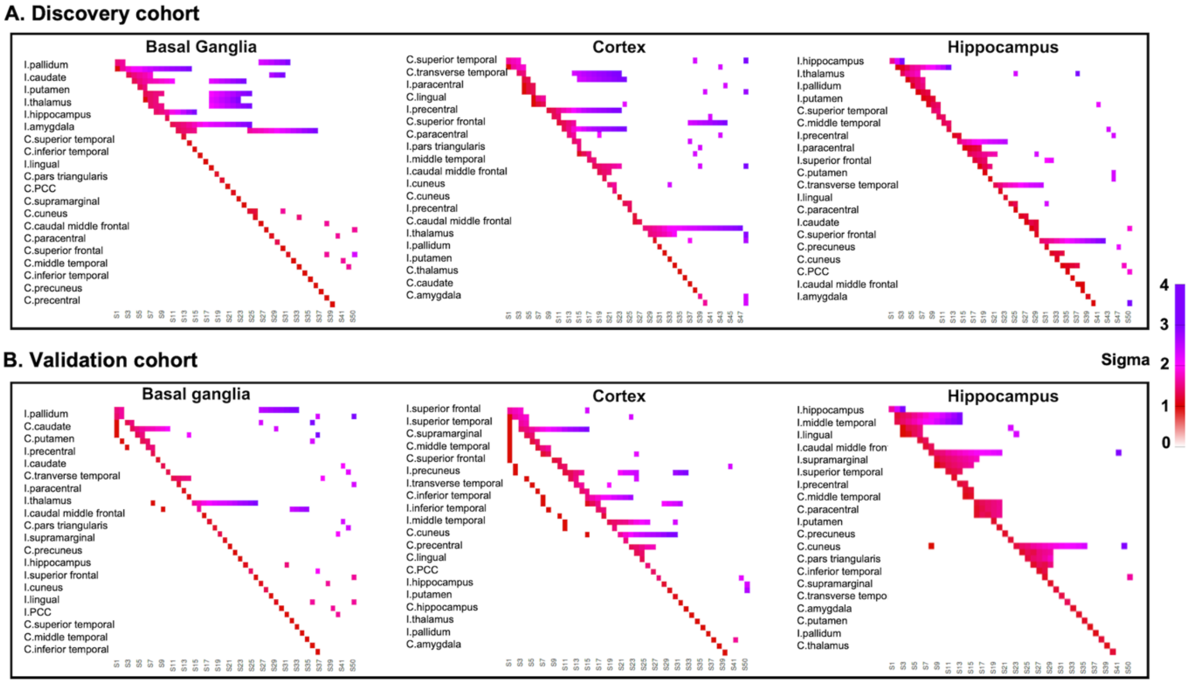


Positional variance diagrams for MRI-based focal epilepsy progression subtypes in the discovery cohort (A) and external validation cohort (B). In both panels, the *y*-axis shows the most likely sequence of atrophy progression, and the *x*-axis shows the position of a given region in the progression sequence, with values ranging from one (first region involved) to the total number of regions. The intensity of each rectangle corresponds to the proportion of Markov Chain Monte Carlo samples of the posterior distribution whereby a certain region of the *y*-axis appears at the respective stage of the *x*-axis. I= ipsilateral; C= contralateral; PCC= posterior cingulate cortex.

**Supplementary Figure 4. Progression subtypes in the IGE external validation cohort.**

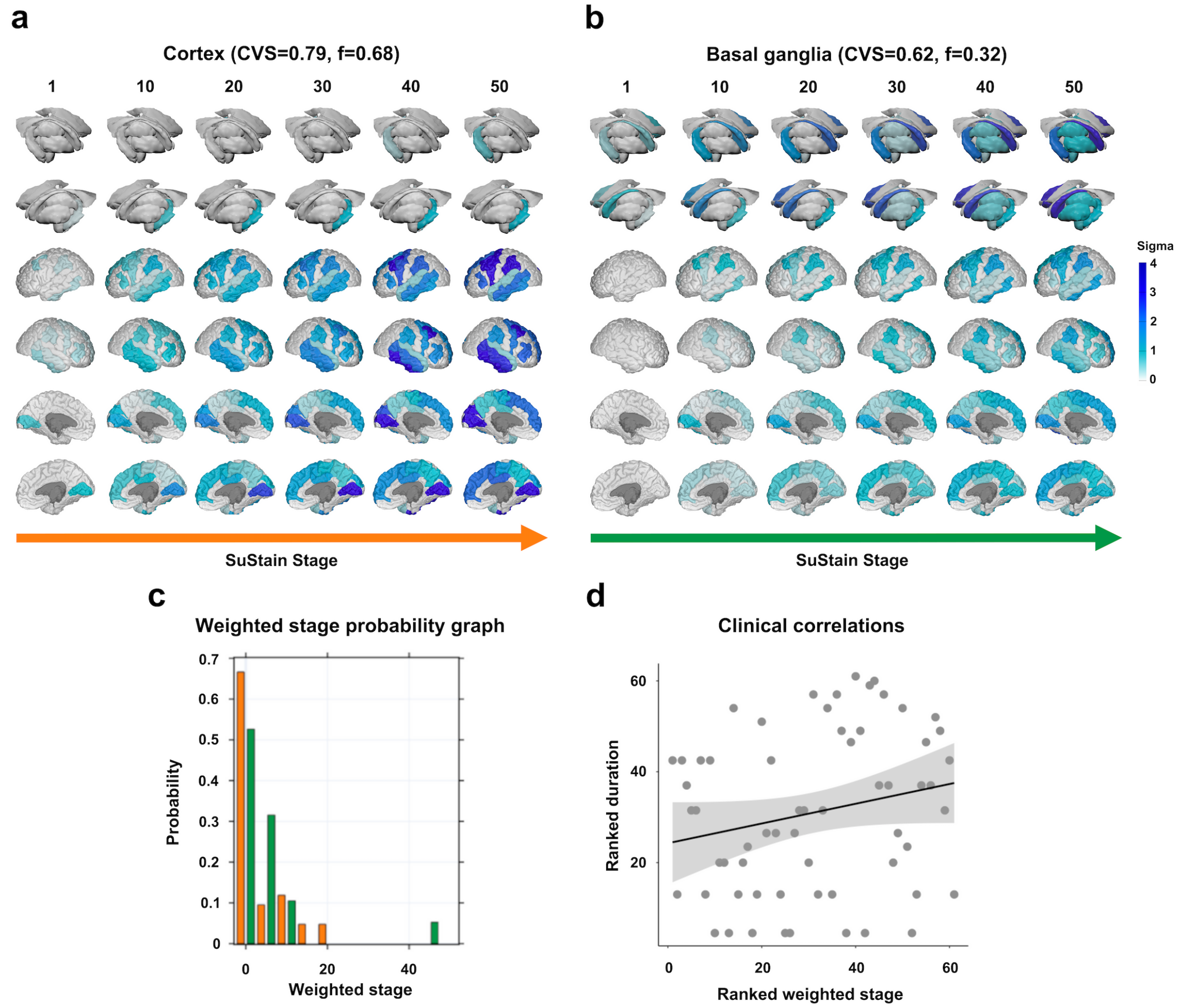


The figure shows the spatiotemporal patterns of progression of grey matter atrophy (*subtypes*; **a.** *cortical*; **b.** *basal ganglia*) identified via SuStaIn in the IGE validation cohort. In panels **a-b**, the colour of each region indicates the severity of grey matter loss; white: unaffected areas; light blue: mildly affected areas (*z*-score=1-2); blue: moderately affected areas (*z*-score=2-3); and dark blue: severely affected areas (*z*-score >3); "CVS": cross-validation similarity. "*f*": proportion of participants assigned to each subtype. Panel **c** shows the probability with which each participant from the IGE discovery cohort was assigned a specific SuStaIn stage (stage ranges: 0.004-53.981). Panel **d** shows the correlation between duration of epilepsy and weighted stage (Spearman's $\rho$) which was not statistically significant; the associated panels show ranked data.
